# Cohort Profile: HUNT4 70+

**DOI:** 10.1101/2025.06.24.25330190

**Authors:** Håvard Kjesbu Skjellegrind, Pernille Thingstad, Linda Gjøra, Marit Kolberg, Grete Kjelvik, Linda Ernstsen, Tone Natland Fagerhaug, Arnulf Langhammer, Steinar Krokstad, Bjørn Olav Åsvold, Marit Næss, Geir Selbæk

## Abstract

HUNT4 70+ is a sub-cohort of persons aged at least 70 years at the 4^th^ survey of the HUNT Study, established to provide data for ageing research.

This population-based sample consists of 9956 individuals from the original HUNT catchment area, included between August 2017 and February 2019. In addition, an urban sample of 1743 persons was included in Trondheim city during October 2018 – June 2019. HUNT4 70+ covers comprehensive aspects of ageing health, including clinical examinations, performance-based tests of physical and cognitive function, questionnaires, and biological samples.

High participation rates among the old and frail were obtained by examination in private homes and nursing homes when needed (15% of the participants). The data can be linked to all national registers in Norway, such as cause of death, prescription, health care utilization, and diagnosis registries.

Data access requires approval from a Norwegian research ethics committee (REC) before application to HUNT Research Centre. Contact HUNT Research Centre for collaboration and more info (ntnu.edu/hunt).

## Why was the cohort set up?

As a part of the Trøndelag Health Study (The HUNT Study), The HUNT4 70+ cohort mainly focuses on physical performance, cognitive function, nutritional status, and oral health in a population-based sample of older adults without exclusion criteria. Other population-based studies have reported disease and symptom burden among older adults attending examination sites, but few studies have reported the continuum from the healthy to those in need of home care or nursing homes.

Previously, dementia has been found in 83% of nursing home residents in the HUNT catchment area [1]. Malnutrition is widespread among older adults [2] and strongly associated with morbidity and mortality [3,4]. Poor oral health has been reported among nursing home residents [5]. Frailty awareness has increased over the last decades, after seminal publications [6,7]. Still, the prevalences of these conditions in the general population were largely unknown. The survey was designed to collect age-related endpoints as a follow-up of the main HUNT cohort and as a baseline for the study of trajectories through old age. Because there are no major cities in the original HUNT catchment area, a supplementary urban cohort was established, HUNT4 Trondheim 70+.

The survey was designed in cooperation with the HUNT ethics committee and approved by the Norwegian Data Protection Authority. Collection and storage of these data are regulated by the “Regulation on population-based health surveys”. Participants were included based on informed consent. Individuals unable to consent were allowed participation after consent by proxy. All were assumed able to refuse participation. The study protocol was approved by the regional research ethics committee before piloting and commencing the study (REK 2016/1880).

### Who is in the cohort?

All adult inhabitants in the HUNT catchment area have been invited to the HUNT surveys (Figure 1). HUNT1 was conducted 1984-86 (participation rate 89.4%), including anthropometric measurements, pulse and blood pressure (BP), as well as questionnaires focusing on hypertension, diabetes, and influence by disease on quality of life, as well as x-ray screening for tuberculosis [8]. HUNT2 (1995-97) (participation rate 69.5%) included extended questionnaires, blood samples, and clinical measurements like spirometry, bone densitometry, and hearing[9]. HUNT3 (2006-08) included 54.1% of the local population [10], and in HUNT4 (2017-19) the participation rate was 54.0% in the original catchment area [11].

**Figure 1:**
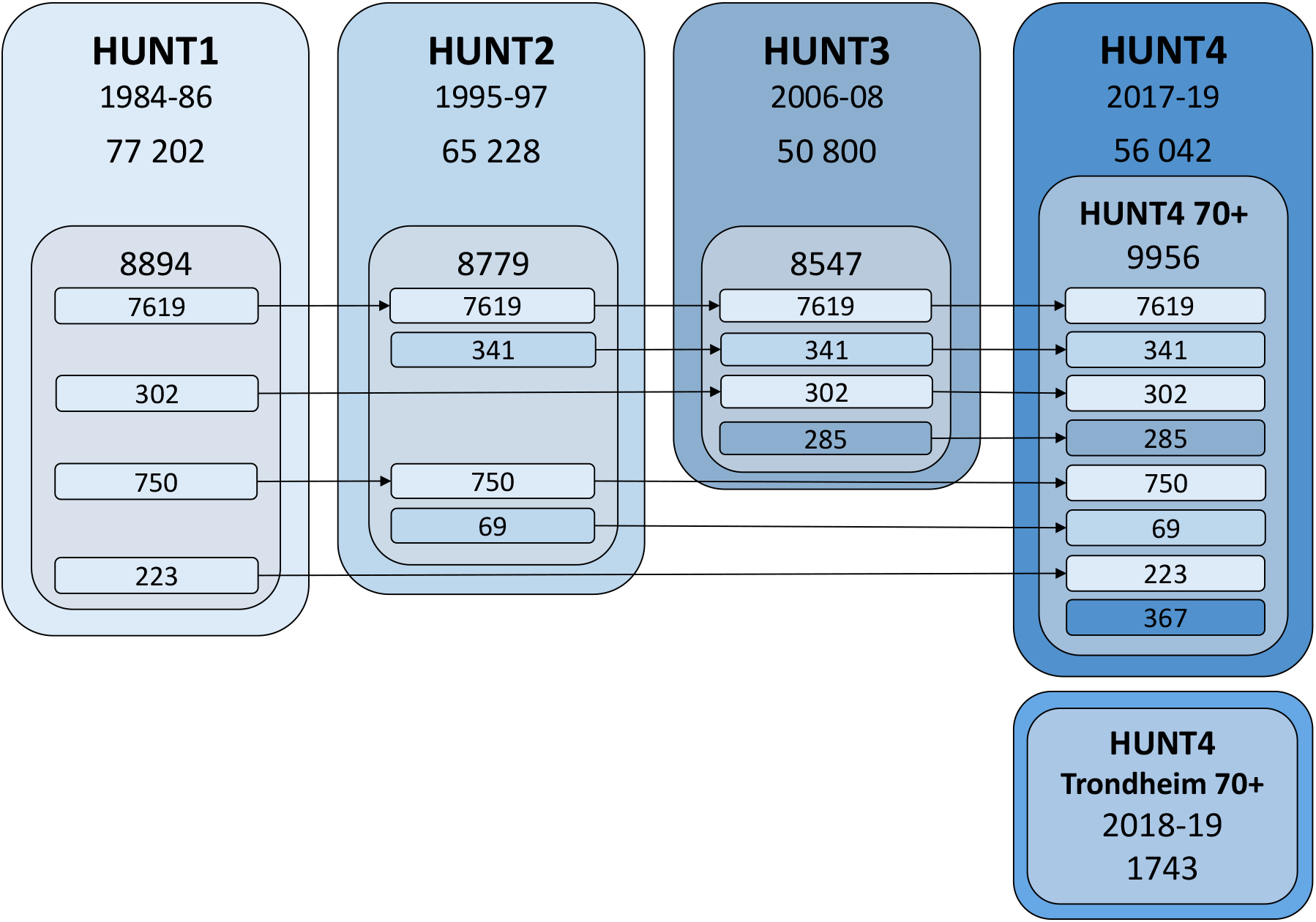
HUNT participation leading up to HUNT4 70+. Figure 1 shows the number of HUNT participants in the different HUNT surveys leading up to participation in HUNT4 70+. The total adult participation in HUNT1 (1984-86) was 77 202, HUNT2 (1995-97) 65 228, HUNT3 (2006-08) 50 800, and HUNT4 (2017-19) 56 042. In the main HUNT4 70+ cohort, 9956 participated. Out of these, 8894 participated in HUNT1, 8779 in HUNT2 and 8547 in HUNT3. HUNT4 Trondheim 70+ included 1743 participants in Trondheim city. These have not been included in previous HUNT surveys.

Out of 19 403 invited older adults aged ≥70 years in the original HUNT catchment area, 12 489 (64.4%) participated in HUNT4 and 9956 (51.3%) were included in the HUNT4 70+ cohort. Examination at home was offered for those not able to attend a field station. Home visits were available to all, by request. Individuals receiving community-based care were given help by their nurses to book a home visit from a HUNT representative. All nursing home residents were invited to participate in an examination in their nursing home. By performing the examinations in several settings, the survey was able to include more older adults and more persons with age-related health conditions.

In addition to the 9956 included in the original HUNT catchment area, 1743 persons aged 70+ out of 5168 invited were included in Trondheim. HUNT4 Trondheim 70+ constitutes an additional, more urban cohort of older adults, with a higher proportion of higher education. As the urban cohort differs in participant characteristics and collected data, it is described separately.

#### Participant profile

To assess representativity of the sample, the point prevalences of common chronic diagnoses were retrieved for 5-year age groups of men and women, for both the included participant sample and the total invited population, from The Norwegian Registry of Primary Health Care. The prevalences of the diagnoses were compared between the participants and the invited population by calculating the relative proportions of diagnosed cases. These comparisons are shown by a heatmap in Figure 2, and in supplementary tables 7 and 8. No systematic difference between the population and the included sample was found. The range of relative proportions spanned from 0.39 to 2.04. For the HUNT4 Trondheim 70+ sample, relative proportions of diagnosed cases are found in supplementary tables 3 and 4.

**Figure 2:**
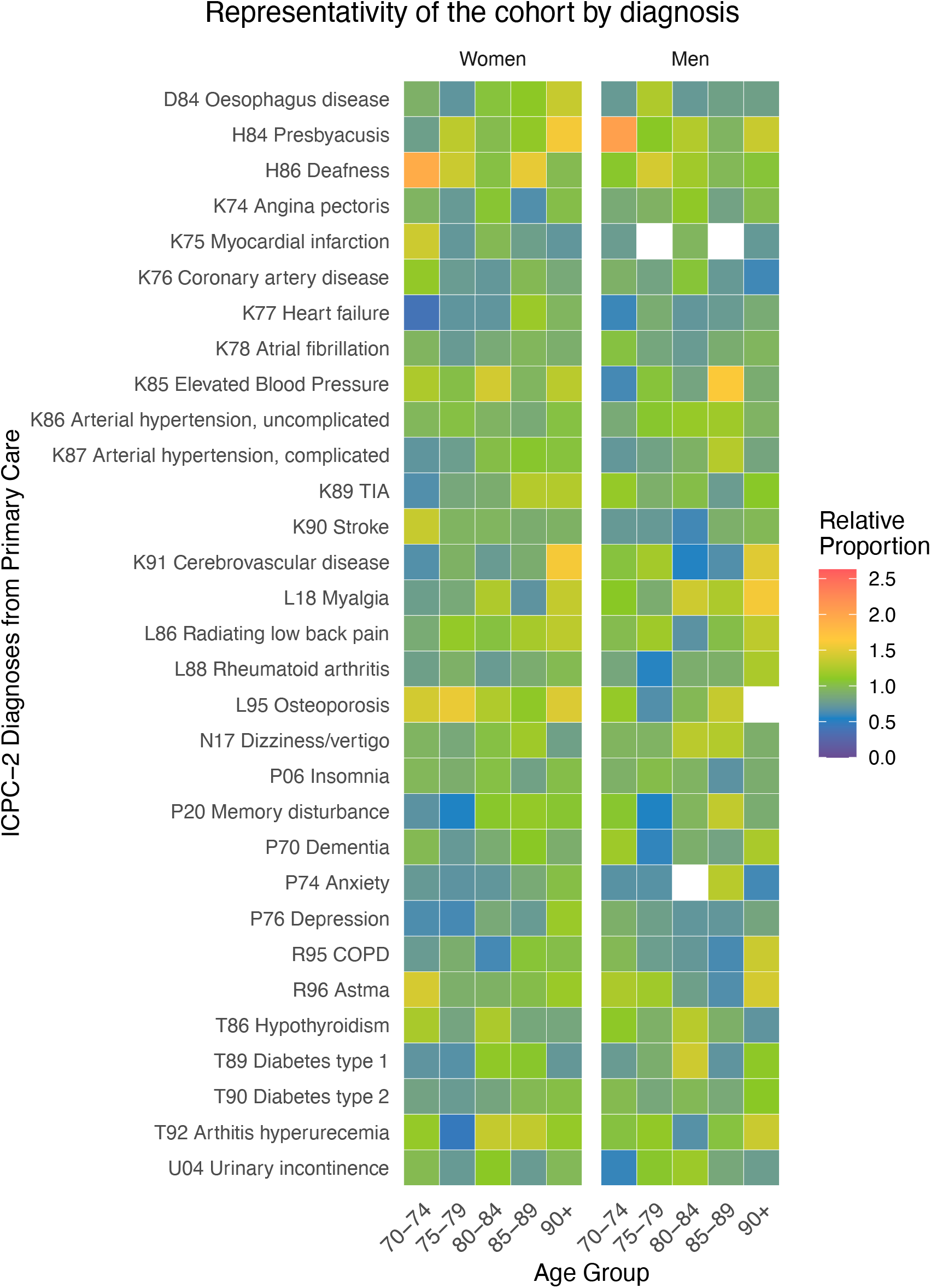
Relative proportions of diagnosis frequency in participants compared to the invited population. Main cohort. Figure 2 shows a heat map representing the relative proportions of diagnosed cases in the cohort compared to the general population, for 31 common conditions, such as cardiovascular, mental and endocrinological conditions. The heat maps are stratified on 5-year age groups and sex and show that for most conditions, the difference between participants in the cohort and the general population is limited. Some age-sex-groups show signs of over-and under-representation, but not with a uniform pattern across ages and sexes.

As the sociodemographic composition of the local and included population may differ from the national population, we have established and published a set of inverse probability weights (IPWs) for participation rate grouped on age, sex and education [12]. IPWs can improve generalisability and precision of prevalence estimates of conditions that are unevenly distributed among these factors, such as dementia, malnutrition, frailty and oral health problems [13–16].

### How often have they been followed up?

Out of the 9956 included participants in the HUNT4 70+ cohort, 9589 have been included in one or several of the former surveys (Figure 1). The HUNT4 70+ cohort was invited to a 4-year follow-up, as a combined effort between the HUNT Ageing in Trøndelag (AiT) and the HUNT COVID projects.

Totally, 5729 individuals from HUNT4 70+ participated, with questionnaires, clinical assessments and blood samples that have been analysed for COVID antibodies. Biomarkers for Alzheimer’s disease (AD) (phosphorylated Tau-217) and axonal damage (Neurofilament Light (NfL)) have been analysed in plasma samples from both HUNT3, HUNT4 and HUNT COVID/AiT. The 4-year follow-up of the HUNT4 Trondheim 70+ cohort included 704 participants with questionnaires and clinical assessments but not biological samples.

Moreover, both the HUNT4 70+ and HUNT4 Trondheim 70+ cohorts can be followed up by linking data to the comprehensive national registers for cause of death, dispensed drugs, diagnosis-specific registries, and diagnoses from health care services, both in primary care and secondary care.

### What has been measured?

#### HUNT4 basic measures

All participants, including those who were examined in their homes or in nursing homes, went through the general HUNT4 examinations as previously described [11]. In brief, BP, pulse, and peripheral oxygen saturation (SpO_2_) were measured three times after a 5-minute rest with 1-minute intervals using an automated BP monitor (GE Carescape V100). At the field stations, body composition, weight, and height were measured using bioelectrical impedance (InBody 770, Cerritos, CA, USA). In private homes and in nursing homes, weight and height were measured using Seca 813 scale and Seca 217 stadiometer (Seca, Germany). Weight and height were measured with light clothes and without shoes. Blood samples were centrifuged within ninety minutes and transported cooled to HUNT biobank for further inhouse processing within 36 hours [17]. Other biomaterials have been collected for parts of the HUNT4 70+ cohort: Urine (44%), saliva (31%), and dried fecal samples (for microbiome) (31%). Genetics is available for 99%. Bioelectrical impedance measurements and biological samples (including blood samples and genetics) were not conducted in HUNT4 Trondheim 70+.

#### Cognitive function

The Montreal Cognitive Assessment (MoCA) assesses executive function, visuospatial abilities, memory, language, attention, abstraction, and orientation, and gives a score ranging 0-30 [18]. The Norwegian translation version 7.1 was used. No additional point was given for low educational attainment.

The CERAD Word List Memory Task (WLMT) tests immediate and delayed recall, which are typically impaired in AD [19]. The task is also sensitive to mild cognitive impairment (MCI) [20]. The WLMT consists of three immediate recall trials of a ten-word list, and then a delayed recall trial. The delayed recall was performed after examination of balance, gait speed, and strength, approximately 5-10 minutes after the immediate recall trials. The WLMT was not applied in cases of MoCA score <22 or zero out of five points in the memory domain of the MoCA.

In nursing homes, for subjects with established moderate to severe cognitive impairment, Severe Impairment Battery-8 [21] was used to assess cognitive function, instead of the MoCA.

The participants were interviewed about subjective cognitive impairments, the development pattern of cognitive impairment, function in activities of daily living, neuropsychiatric symptoms, former examinations on cognitive function, and family history of cognitive impairments.

In nursing homes, a caregiver provided information on symptoms and daily function. Neuropsychiatric symptoms were assessed using the nursing home version of the Neuropsychiatric Inventory (NPI-NH)[22,23]. Activities of daily living (ADL) were assessed using the Physical Self-Maintenance Scale (PSMS)[24].

#### Proxy interview

Participants were asked for permission to contact a next of kin for supplementary information on their cognitive health if at least one of these criteria were met:

- MoCA score below a threshold (70-80 years: <22/30; 80-90 years: <21; 90+: <20)
- WLMT delayed recall score below threshold (70-80 years: <4/10 words; 80-90 years: <3; 90+: <2)
- Self-reported subjective cognitive impairment: Answering yes to EITHER “Has your memory become significantly worse in the last five years?”, OR “Have other cognitive functions (like orientation ability or language) become worse in the last five years?”, AND “Do these changes worry you?”.

When possible, the proxy was chosen by the participant and was in most cases the spouse or a daughter/son. For participants with consent by proxy, that next of kin was interviewed. All interviews were conducted by telephone and included the following scales:

- Neuropsychiatric Inventory Questionnaire (NPI-Q) [25]
- Instrumental activities in daily living (I-ADL) and physical ADL (P-ADL) [24]

The interview also included questions on symptom debut, symptom development, dementia diagnosis, use of municipal and informal care and family history of dementia.

#### Clinical dementia rating (CDR)

CDR was scored by the examiner in cooperation with the nursing staff in nursing homes and based on the proxy interviews [26].

#### Dementia and MCI diagnosis

All available information from questionnaires, interviews, cognitive tests and proxy interviews, were used to establish consensus-based diagnoses of MCI, dementia and dementia subtypes, applying DSM-5 diagnostic criteria [American Psychiatric Association, 15,27].

#### Physical function

The Short Physical Performance Battery (SPPB) is a performance-based test of physical function including three timed sub-tests: 4-meter gait speed, five times sit-to-stand at maximum speed and three standing balance positions [28]. Each subtest is scored 1-4 points or zero points if the person is unable to perform the subtask. The Norwegian translation of the original test protocol and score sheets were used.

One-leg balance was performed both with open and closed eyes, as a timed test for balancing on the preferred one leg and registered as 0-30 seconds or not able. The time was stopped when moving the standing leg or touching down the elevated leg.

Grip strength was measured by using the Jamar Plus+ Digital Hand Dynamometer® (Performance Health, Warrenwille, USA). Three trials were performed consecutively without resting pauses for each hand with the person in sitting position, the arm hanging free by the body, the shoulder adducted and neutrally rotated, the elbow held in 90° and the wrist in neutral position.

#### 24-hour movement behaviour

Two AX3 accelerometers (Axivity, Newcastle, UK), one placed on the thigh and one on the lower back were worn for seven days [29]. Validated machine learning models have been used to predict the amount of time of different types of physical activity [30] and duration of sleep [31].

#### Nutritional assessment

In addition to height and weight, the mid-upper arm circumference (MUAC) was measured for nutritional assessment. MUAC was measured twice on the right upper arm at the midpoint between acromion and olecranon. Interview and questionnaire questions included unintentional weight loss, food intake, loss of appetite, problems with chewing food and oral pain, enabling evaluation of malnutrition [3].

#### Oral health

The Revised Oral Assessment Guide (ROAG) is a standardized assessment tool for nursing staff to examine and detect oral health problems in older adults [32]. ROAG-Jönköping (ROAG-J) is a modified ROAG and includes assessment of voice, swallowing, lips, oral mucosa, tongue, saliva, gums, teeth and dentures. Additionally, information about saliva, number of teeth, mobility of teeth, root remnants and dentures were registered. The lower end of a toothbrush was used to test buccal mucosal surface sliding friction. In the main cohort, ROAG-J was performed at nursing homes and home visits only. In HUNT4 Trondheim 70+, ROAG-J was performed for all participants.

The number of included participants with data on different measures are shown in table 2. The distribution of selected key variables are shown in Figure 3 for the HUNT4 70+ cohort and in supplementary Figure S1 for HUNT4 Trondheim 70+.

**Table 1:** Participant characteristics.

|  | HUNT4 70+<br>(N=9956) | Trondheim 70+<br>(N=1743) |
| --- | --- | --- |
| **Sex** |  |  |
| Female | 5420 (54.4%) | 1008 (57.8%) |
| Male | 4536 (45.6%) | 735 (42.2%) |
| **Age stratum** |  |  |
| 70-79 | 6712 (67.4%) | 1128 (64.7%) |
| 80-89 | 2579 (25.9%) | 450 (25.8%) |
| 90+ | 665 (6.7%) | 165 (9.5%) |
| **Education** |  |  |
| Primary | 3173 (31.9%) | 193 (11.1%) |
| Secondary | 4217 (42.4%) | 557 (32.0%) |
| College/University | 2362 (23.7%) | 707 (40.6%) |
| **Smoking Status** |  |  |
| Never | 3548 (35.6%) | 585 (33.6%) |
| Former | 4653 (46.7%) | 342 (19.6%) |
| Current | 578 (5.8%) | 59 (3.4%) |
| Occasional | 28 (0.3%) | 20 (1.1%) |
| Former Occasional | 419 (4.2%) | 434 (24.9%) |
| **ApoE ε4 carrier** |  |  |
| No | 6875 (69.1%) | - |
| Yes | 2970 (29.8%) | - |
| **BMI (kg/m<sup>2</sup>)** | 27.2 (4.4) | 26.6 (4.4) |
| **Body fat (%)** | 33.6 (8.5) | - |
| **MUAC (cm)** | 29.9 (3.6) | 29.3 (3.9) |
| **Systolic BP (mmHg)** | 139.4 (20.3) | 138.9 (21.0) |
| **Grip Strength (kg)** | 31.9 (11.5) | 29.5 (11.5) |
| **SPPB** | 11 [8, 12] | 11 [8, 12] |
| **ROAG-J<sup>a</sup>** |  |  |
| Normal | - | 674 (38.7%) |
| ≥1 Moderate Problem | - | 721 (41.4%) |
| ≥1 Severe Problem | - | 167 (9.6%) |
| **MoCA** | 24 [20, 26] | 24 [21, 26] |
| **CERAD 10-word recall** | 5 [4, 6] | 5 [4, 7] |
| **Dementia assessment** |  |  |
| No cognitive impairment | 4822 (48.4%) | 796 (45.7%) |
| aMCI | 2920 (29.3%) | 439 (25.2%) |
| naMCI | 497 (5.0%) | 121 (6.9%) |
| Dementia | 1531 (15.4%) | 363 (20.8%) |
| Unclassified | 186 (1.9%) | 24 (1.4%) |

Continuous variables are presented as mean (SD), and discrete scores as median [IQR]. Percentages are of total N in column. Missing is not shown. BMI: Body Mass Index, BP: Blood Pressure. MUAC: Middle Upper Arm Circumference, SPPB: Short Physical Performance Battery, ROAG-J: Revised Oral Assessment Guide – Jönköping, CERAD: Consortium to Establish a Registry for Alzheimer's Disease. aMCI: amnesic mild cognitive impairment (MCI), naMCI: non-amnesic MCI. <sup>a</sup>ROAG-J was only assessed at home visits (n=699) and nursing homes (n=426) in the main cohort, see suppl. table 1 and 5 for setting-specific participant characteristics.

**Table 2:** Data availability.

| **Data** | **HUNT4 70+**<br>**(N=9956)** | **Trondheim 70+**<br>**(N=1743)** |
| --- | --- | --- |
| **Questionnaire** |  |  |
| Education | 9752 ( 98.0%) | 1457 ( 83.6%) |
| Smoking status | 9226 ( 92.7%) | 1440 ( 82.6%) |
| Weight loss | 8447 ( 84.8%) | 1329 ( 76.2%) |
| Exhaustion | 7826 ( 78.6%) | 1380 ( 79.2%) |
| HADS | 7554 ( 75.9%) | 1351 ( 77.5%) |
| BMI | 9458 ( 95.0%) | 1576 ( 90.4%) |
| Body Composition | 7565 ( 76.0%) | - |
| MUAC | 8682 ( 87.2%) | 1691 ( 97.0%) |
| Systolic BP | 9791 ( 98.3%) | 1693 ( 97.1%) |
| Gait Speed | 9142 ( 91.8%) | 1589 ( 91.2%) |
| Grip Strength | 9337 ( 93.8%) | 1681 ( 96.4%) |
| SPPB | 9067 ( 91.1%) | 1647 ( 94.5%) |
| 1-week Accelerometer recording | 4863 ( 48.8%) | 1184 ( 67.9%) |
| ROAG-J | 1125 ( 11.3%) | 1562 ( 89.6%) |
| MoCA | 8908 ( 89.5%) | 1480 ( 84.9%) |
| CERAD 10-word recall | 5439 ( 54.6%) | 1019 ( 58.5%) |
| SIB-8 | 356 ( 3.6%) | 138 ( 7.9%) |
| Dementia assessment | 9775 ( 98.2%) | 1721 ( 98.7%) |
| **Lab data (examples)** |  |  |
| Cholesterol | 9539 ( 95.8%) | - |
| pTau-217 | 8948 ( 89.9%) | - |
| NfL | 8789 ( 88.3%) | - |
| Genetics | 9889 ( 99.3%) | - |
| **Bio-samples** |  |  |
| Serum | 9468 ( 95.1%) | - |
| Plasma | 9063 ( 91.0%) | - |
| Urine | 4391 ( 44.1%) | - |
| Faeces | 3051 ( 30.6%) | - |
| Saliva | 3044 ( 30.6%) | - |
| HUNT3 Serum | 8385 ( 84.2%) | - |
| HUNT3 Plasma | 7506 ( 75.4%) | - |
| HUNT2 Serum | 8720 ( 87.6%) | - |

HADS: Hospital Anxiety and Depression Scale, BMI: Body Mass Index, MUAC: Middle Upper Arm Circumference, SPPB: Short Physical Performance Battery, ROAG-J: Revised Oral Assessment Guide – Jönköping, MoCA: Montreal Cognitive Assessment, CERAD: Consortium to Establish a Registry for Alzheimer's Disease. SIB-8: Severe Impairment Battery-8. pTau-217: plasma phosphorylated Tau-217, NfL, plasma neurofilament light. See supplemental table 2 and 6 for data availability stratified on data collection settings.

**Figure 3:**
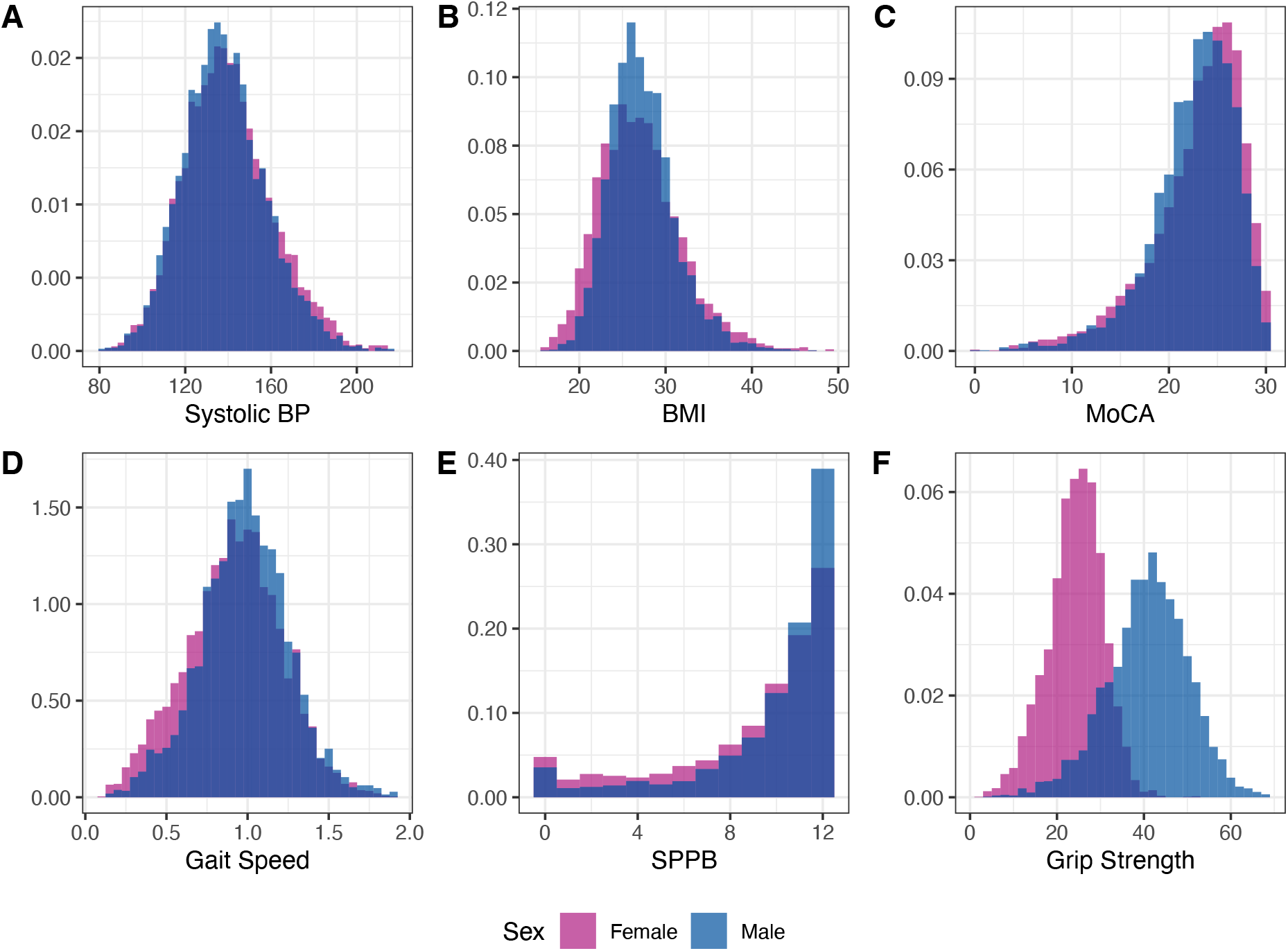
Distribution of key variables. A Systolic blood pressure (mmHg), B Body mass index (kg/m2), C Montreal Cognitive Assessment, D Gait speed (m/s), E Short Physical Performance Battery, F Grip Strength. The figure shows histograms representing the distribution of values for Systolic blood pressure, body mass index (BMI), Montreal Cognitive Assessment (MoCa), Gait Speed, Short Physical Performance Battery (SPPB), and Grip Strength. The histograms are stratified by sex. Blood pressure is normally distributed with a mean of 138 mmHg for men and 140 mmHg for women. BMI is close to normally distributed, with a mean of 27 kg/m^2^ for both women and men. MoCA is left skewed with a mean of 23 for both women and men. Gait speed is normally distributed, with a mean of 0.98 m/s for men and 0.91 m/s for women. SPPB is heavily left skewed, showing an obvious roof effect, with a median of 11 for men and 10 for women, out of max 12. Grip strength is close to normally distributed but contrary to the other variables shown, the difference between women and men is substantial, with a mean of 24 kg for women and 40 kg for men.

### What has it found?

The HUNT4 70+ cohort was used to estimate current and future prevalence of dementia and MCI in Norway [15]. Standardized dementia prevalence ranged from 5.6% at 70-74 years, increasing to 48.1% at 90+ years. MCI was found in 33-36% in all ages ≥70. Moreover, we found that only 35.6% of those with a dementia condition were diagnosed in the health care system [33]. The four-year follow-up was used to estimate a yearly dementia incidence of 4.4% [34].The cohort has been used to establish norms for the MoCA [35] and the WLMT [36]. We have also described the variation of physical performance with age and education [37]. A HUNT Frailty Index (HUNT-FI) has been constructed and, together with the Fried phenotype, used to establish the prevalence of frailty in the Norwegian 70+ population [38]. Physical performance was found proportionally impaired with cognitive impairment, also with MCI [39]. The prevalence of malnutrition in the population aged 70+ was estimated to 14.3% [13] and the prevalence of oral health problems 56.1% [16].

The cohort is well suited to study risk factors for dementia. Using BP from all HUNT surveys, we have found that, on average, BP increases with increasing age in those that maintain cognitive function, while in those developing dementia, average BP declined in advanced age. Still, ten years before dementia diagnosis, average BP was the same [40]. A combination of a low BP trajectory and high physical activity was associated with the lowest dementia risk [41]. A study on hearing loss identified in HUNT2 (1995-97) found this associated to dementia risk 22 years later [42].

An overview of published papers is found in the supplement.

### What are the main strengths and weaknesses?

The HUNT4 70+ survey comprises a comprehensive dataset covering important aspects of ageing health for a representative sample, including the oldest and frailest part of the population. In total, 51.4% of the entire population in the original HUNT catchment area was included. This, combined with having both clinical data and biological samples for most participants over several decades is a major strength. In addition, the data may be linked to a wide range of national and local health registries, including cause of death, cancer, primary and secondary care diagnoses, socioeconomic indicators, drug prescriptions, and health care utilization. The very long follow-up time of 33 years from HUNT1 and 22 years from the first blood samples (HUNT2) allows studies with low risk of reverse causation, and the comprehensiveness allows analyses with low levels of residual confounding. The addition of HUNT4 Trondheim 70+ resolves the previous lack of an urban population.

Missing questionnaire data is a potential weakness, especially for individuals with severe cognitive impairments. However, some data, such as education, may be found in questionnaires from previous HUNT surveys. For risk assessment studies, the competing risk of death may be a weakness. Cause of death data may be acquired through the Norwegian Cause of Death register. The participation rate of approximately 50% may be considered low for some purposes. We find that using weighed analyses may compensate this well [14].

### Can I get hold of the data? Where can I find out more?

The HUNT4 70+ dataset was collected to provide the public with more extensive epidemiological research on ageing and age-related health. Thus, we invite researchers to apply to HUNT Research Centre to obtain data for projects. Data access requires approval from a Norwegian Research Ethics Committee (REC) following an application submitted by a researcher affiliated with a Norwegian research institution. We advise contacting HUNT Research Centre to establish relevant collaborations. A complete catalogue of available variables is found at ntnu.edu/hunt/data.

## Supporting information

Supplement for Cohort Profile HUNT4 70+

## Data Availability

Data access requires approval from a Norwegian Research Ethics Committee (REC) following an application submitted by a researcher affiliated with a Norwegian research institution. We advise contacting HUNT Research Centre to establish relevant collaborations. A complete catalogue of available variables is found at ntnu.edu/hunt/data.

https://www.ntnu.no/hunt

## Ethics approval

The data collection in the HUNT Study is legally regulated and not subject to external approval requirements from data security, privacy or research ethics authorities. This cohort article was approved by the Mid-Norway Regional Committee for Medical and Health Research Ethics (REK 69134).

## Acknowledgements

The HUNT4 70+ survey has received funding from the Norwegian National Centre for Ageing and Health and Center for Oral Health Services and Research (TkMidt), Trondheim, in addition to funding of HUNT4 by the Ministry of Health and Care Services, Central Norway Regional Health Authority, Trøndelag county council and NTNU. All municipalities have contributed with staff and field station offices. ROAG was used in HUNT4 70+ with permission from Pia Andersson. The ROAG-J and related guidance manual was used as part of the training of the examiners with permission by Eva Herremo and Charlott Karlsson, Folktandvården, Region Jönköpings län and Senior Alert, Qulturum, Region Jönköpings län. ROAG-J was translated to Norwegian by Center for Oral Health Services and Research (TkMidt) to be used in HUNT4 70+. Data from the Norwegian Registry for Primary Health Care has been used in this publication. The interpretation and reporting of these data are the sole responsibility of the authors, and no endorsement by the Dep. of Health Registries is intended nor should be inferred.

All data analysis and visualization in this article were performed using R version 4.4.1 “Race for Your Life” in RStudio Version 2024.04.2+764 “Chocolate Cosmos”. Tables were made using the table1 and flextable packages, and graphics were made using ggplot2.

## Author contributions

Design and execution of the data collection: all authors. Management of the data collection: PT, LG and HKS. Drafting the manuscript: HKS. Data analysis and visualization: HKS. Review and final approval of the manuscript: all authors.

## Use of artificial intelligence (AI) tools

The manuscript is entirely human written, and solely a product by the authors. Yale Clarity (GPT-4o) has been used for debugging of R code written to prepare tables and figures. AI has not been used for interpretation, citations or other content generation.

## Supplementary data

Supplementary data are available online.

