## Supplement for Cohort Profile HUNT4 70+ for "Cohort Profile: HUNT4 70+"

**HUNT4 Trondheim 70+ – an urban extension of the HUNT4 70+ cohort**

The HUNT4 70+ survey was extended beyond the original HUNT catchment area by including parts of Trondheim city. This was set up to include an urban population. This survey part was performed October 2018 to June 2019. Participants from one out of four districts in Trondheim city were included using the same questionnaire and examinations as in the HUNT4 70+ catchment area, except no biological samples were collected and bioelectrical impedance measurements were not performed. Participants were included by field station attendance (73.9%), home visit (11.3%) and in nursing homes (14.8%). Out of 5168 invited 1743 (33.6 %) participated.

*Methods:*

The data collection was largely performed according to the protocol used in the main HUNT4 70+ cohort, with some modifications:

Weight and height were measured manually by Seca scale and Seca stadiometer.

The oral health examination (ROAG-J) was performed for all participants, both in the field station, at home visits and nursing homes. In the main HUNT4 70+ cohort the ROAG-J was performed only for participants in nursing homes and home examinations, due to capacity reasons.

The participants had the opportunity to fill the self-report questionnaires while at the field station, opposed to the main cohort who did this at home before and after the clinical examinations. Printed questionnaires were available for all, both in Trondheim and Nord-Trøndelag.

*Strengths and weaknesses:*

We have included 1485 community-dwelling older adults in a major Norwegian city, and thus added an urban population to the main HUNT4 70+ cohort. We have shown previously that differences in educational attainment in this region had impact on the prevalence of dementia (Gjøra et al., 2023). However, by use of inverse probability weighting analysis, we found similar prevalences for dementia in Trondheim and the original HUNT catchment area (Nord-Trøndelag).

The nursing home part of the survey is up-sampled compared to the general population. This was not intentional but merely a result of a higher participation rate among nursing home residents compared to the home-dwelling older population. This sample may be used in conjunction with the community-dwelling participants (preferably with weighting correcting for the up-sampling) or in combination with the nursing home sample in the main cohort to study residents in long term nursing facilities. The Trondheim 70+ sample show similar distributions of key variables (Supplemental figure 1) as in the main cohort, also here with major sex-differences in muscular strength.

The HUNT4 Trondheim 70+ survey constitute an important data set especially for studies using the ROAG-J, as this examination was performed at all settings and thus for nearly all the participants.

The individuals surveyed in HUNT4 Trondheim 70+ have not participated in the previous surveys of The HUNT Study. Thus, background data lack for this part of the cohort. Also, this sample does not have bio-samples, nor genetics. The Trondheim population also has higher level of education compared to the Nord-Trøndelag population. Missing data for those unable to fill the questionnaires is pronounced especially among those included in home visits and nursing homes. Without data from previous surveys, linking to Statistics Norway may be necessary to include key variables such as educational attainment.

Supplemental table 1 – Participant characteristics
HUNT4 Trondheim 70+

| | Field station (N=1287) | Home visit (N=198) | Nursing home (N=258) | Overall (N=1743) |
|---|---|---|---|---|
| **Sex** | | | | |
| Female | 690 (53.6%) | 131 (66.2%) | 187 (72.5%) | 1008 (57.8%) |
| Male | 597 (46.4%) | 67 (33.8%) | 71 (27.5%) | 735 (42.2%) |
| **Age stratum** | | | | |
| 70-79 | 1023 (79.5%) | 57 (28.8%) | 48 (18.6%) | 1128 (64.7%) |
| 80-89 | 247 (19.2%) | 100 (50.5%) | 103 (39.9%) | 450 (25.8%) |
| 90+ | 17 (1.3%) | 41 (20.7%) | 107 (41.5%) | 165 (9.5%) |
| **Education** | | | | |
| Primary | 107 (8.3%) | 38 (19.2%) | 48 (18.6%) | 193 (11.1%) |
| Secondary | 474 (36.8%) | 44 (22.2%) | 39 (15.1%) | 557 (32.0%) |
| College/university | 653 (50.7%) | 29 (14.6%) | 25 (9.7%) | 707 (40.6%) |
| **Smoking Status** | | | | |
| Never | 490 (38.1%) | 46 (23.2%) | 49 (19.0%) | 585 (33.6%) |
| Former | 286 (22.2%) | 22 (11.1%) | 34 (13.2%) | 342 (19.6%) |
| Current | 56 (4.4%) | 3 (1.5%) | 0 (0%) | 59 (3.4%) |
| Occasional | 16 (1.2%) | 1 (0.5%) | 3 (1.2%) | 20 (1.1%) |
| Former Occasional | 369 (28.7%) | 33 (16.7%) | 32 (12.4%) | 434 (24.9%) |
| **BMI (kg/m2)** | 26.6 (4.1) | 27.0 (5.4) | 26.1 (5.3) | 26.6 (4.4) |
| **MUAC (cm)** | 29.7 (3.6) | 28.6 (4.7) | 27.5 (4.1) | 29.3 (3.9) |
| **Systolic BP (mmHg)** | 139.6 (19.8) | 137.1 (22.6) | 136.2 (25.9) | 138.9 (21.0) |
| **Grip Strength (kg)** | 32.8 (10.2) | 23.2 (8.9) | 15.7 (7.4) | 29.5 (11.5) |
| **SPPB** | 11 [10, 12] | 5 [2, 9] | 1 [0, 3] | 11 [8, 12] |
| **ROAG-J** | | | | |
| Normal | 587 (45.6%) | 57 (28.8%) | 30 (11.6%) | 674 (38.7%) |
| ≥1 Moderate Problem | 512 (39.8%) | 89 (44.9%) | 120 (46.5%) | 721 (41.4%) |
| ≥1 Severe Problem | 93 (7.2%) | 16 (8.1%) | 58 (22.5%) | 167 (9.6%) |
| **MoCA** | 25 [22, 27] | 20 [16, 23] | 13 [8, 15] | 24 [21, 26] |
| **CERAD 10-word recall** | 5 [4, 7] | 4 [3, 6] | - | 5 [4, 7] |
| **Dementia assessment** | | | | |
| No cognitive impairment | 737 (57.3%) | 54 (27.3%) | 5 (1.9%) | 796 (45.7%) |
| aMCI | 377 (29.3%) | 59 (29.8%) | 3 (1.2%) | 439 (25.2%) |
| naMCI | 93 (7.2%) | 16 (8.1%) | 12 (4.7%) | 121 (6.9%) |
| Dementia | 76 (5.9%) | 50 (25.3%) | 237 (91.9%) | 363 (20.8%) |
| Unclassified | 4 (0.3%) | 19 (9.6%) | 1 (0.4%) | 24 (1.4%) |

Continuous variables are presented as mean (SD), and discrete scores as median [IQR]. Percentages are of total N in column. Missing is not shown. BMI: Body Mass Index, MUAC: Middle Upper Arm Circumference, SPPB: Short Physical Performance Battery, ROAG-J: Revised Oral Assessment Guide – Jönköping, CERAD: Consortium to Establish a Registry for Alzheimer's Disease. aMCI: amnestic mild cognitive impairment (MCI), naMCI: non-amnestic MCI.

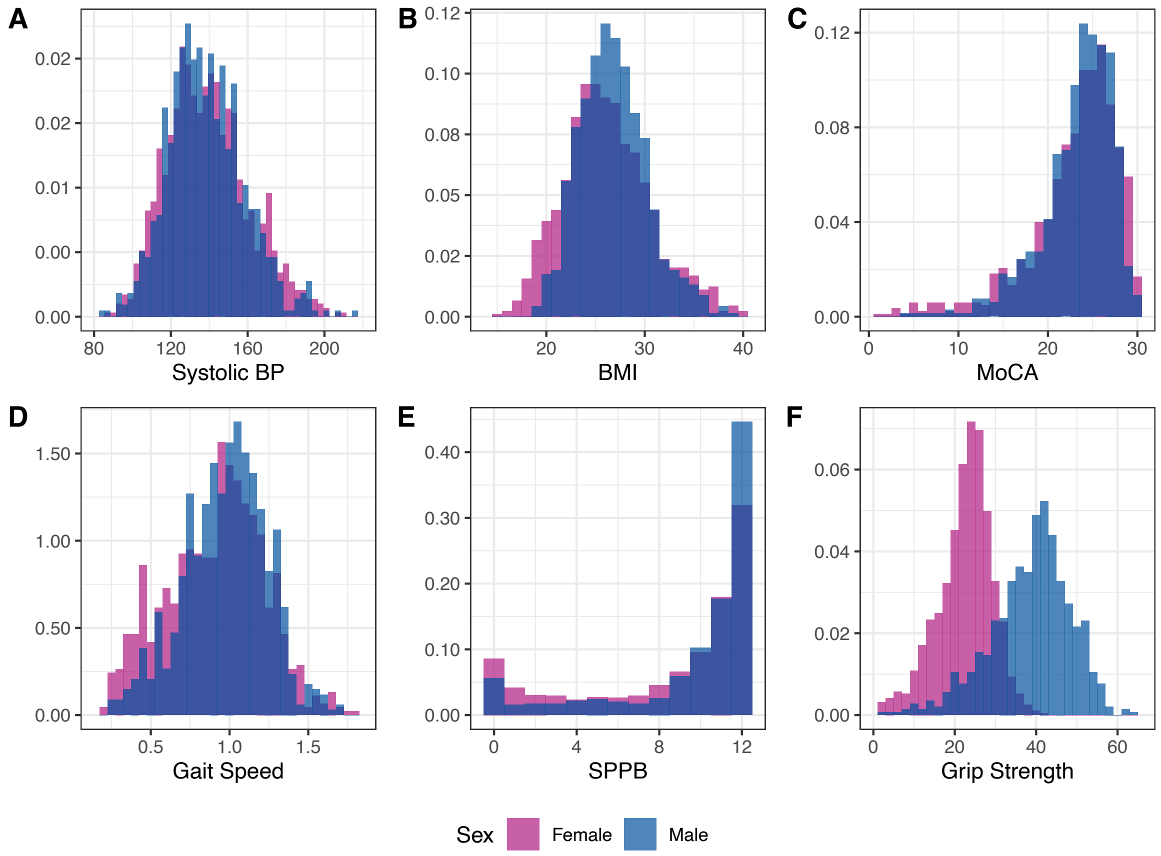


*Supplemental figure S1 - distribution of key variables*
*A Systolic blood pressure (mmHg), B Body mass index (kg/m²), C Montreal Cognitive Assessment, D Gait speed (m/s), E Short Physical Performance Battery, F Grip Strength (kg).*

*Assessment of representativity*
To assess representativity of the included sample, the included persons were compared to the local population in the same age groups, such as for the main cohort, by diagnose frequency retrieved from the The Norwegian Registry of Primary Health Care. As the registry do not release numbers for groups less than five persons, a high number of combinations of 5-year age group, sex and diagnose have missing values in this smaller sample of individuals. Such missingness was especially pronounced for the oldest men. The relative proportions of diagnose cases (with 95% confidence intervals) for women and men in 5-year age groups are included in supplementary table 3 and 4 and illustrated by a heatmap in figure S2. The range of relative proportions spanned from 0.46 to 3.24.

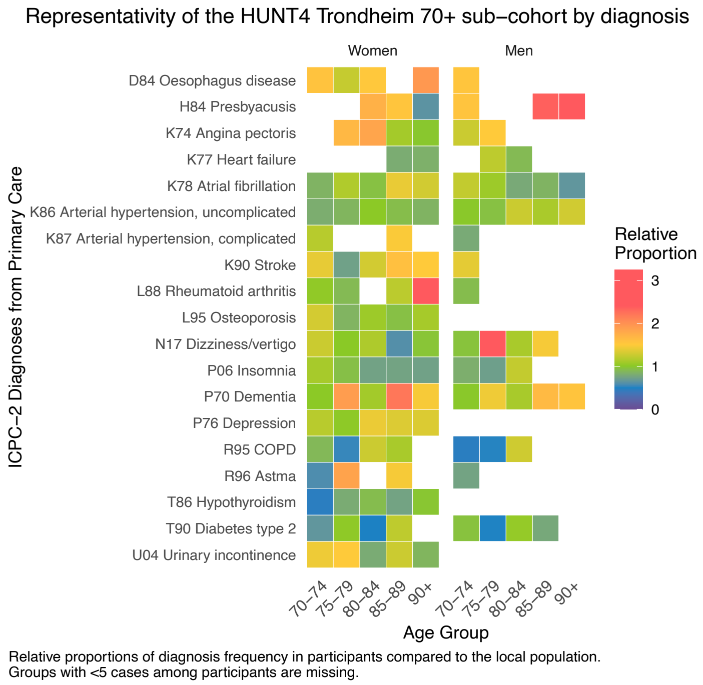


*Supplemental figure S2*

Supplemental table 2 – Data availability HUNT4 Trondheim 70+

| Data | Field station (N=1287) | Home visit (N=198) | Nursing home (N=258) | Total (N=1743) |
|---|---|---|---|---|
| **Questionnaire** | | | | |
| Education | 1234 ( 95.9 %) | 111 ( 56.1 %) | 112 ( 43.4 %) | 1457 ( 83.6 %) |
| Smoking status | 1217 ( 94.6 %) | 105 ( 53 %) | 118 ( 45.7 %) | 1440 ( 82.6 %) |
| Weight loss | 1141 ( 88.7 %) | 105 ( 53 %) | 83 ( 32.2 %) | 1329 ( 76.2 %) |
| Exhaustion | 1188 ( 92.3 %) | 105 ( 53 %) | 87 ( 33.7 %) | 1380 ( 79.2 %) |
| HADS | 1186 ( 92.2 %) | 98 ( 49.5 %) | 67 ( 26 %) | 1351 ( 77.5 %) |
| **Clinical examinations** | | | | |
| BMI | 1276 ( 99.1 %) | 167 ( 84.3 %) | 133 ( 51.6 %) | 1576 ( 90.4 %) |
| MUAC | 1283 ( 99.7 %) | 186 ( 93.9 %) | 222 ( 86 %) | 1691 ( 97 %) |
| Systolic BP | 1282 ( 99.6 %) | 190 ( 96 %) | 221 ( 85.7 %) | 1693 ( 97.1 %) |
| Gait Speed | 1282 ( 99.6 %) | 151 ( 76.3 %) | 156 ( 60.5 %) | 1589 ( 91.2 %) |
| Grip Strength | 1281 ( 99.5 %) | 185 ( 93.4 %) | 215 ( 83.3 %) | 1681 ( 96.4 %) |
| SPPB | 1238 ( 96.2 %) | 172 ( 86.9 %) | 237 ( 91.9 %) | 1647 ( 94.5 %) |
| 1-week Accelerometer recording | 1028 ( 79.9 %) | 85 ( 42.9 %) | 71 ( 27.5 %) | 1184 ( 67.9 %) |
| ROAG-J | 1192 ( 92.6 %) | 162 ( 81.8 %) | 208 ( 80.6 %) | 1562 ( 89.6 %) |
| MoCA | 1267 ( 98.4 %) | 156 ( 78.8 %) | 57 ( 22.1 %) | 1480 ( 84.9 %) |
| CERAD 10-word recall | 964 ( 74.9 %) | 55 ( 27.8 %) | - | 1019 ( 58.5 %) |
| SIB-8 | - | - | 138 ( 53.5 %) | 138 ( 7.9 %) |
| Dementia assessment | 1284 ( 99.8 %) | 179 ( 90.4 %) | 258 ( 100 %) | 1721 ( 98.7 %) |

HADS: Hospital Anxiety and Depression Scale, BMI: Body Mass Index, MUAC: Middle Upper Arm Circumference, SPPB: Short Physical Performance Battery, ROAG-J: Revised Oral Assessment Guide – Jönköping, MoCA: Montreal Cognitive Assessment, CERAD: Consortium to Establish a Registry for Alzheimer's Disease. SIB-8: Severe Impairment Battery-8.

**What has been found?**

*Published results:*

The HUNT4 Trondheim 70+ data has been used separately for some purposes, such as norms for cognitive tests, and prevalences based on the ROAG-J. Education- and age-adjusted norms for the Norwegian version of Mini-Mental State Examination (MMSE-NR3) were estimated based on a subsample from HUNT4 Trondheim 70+ (Engedal et al., 2023). Also, norms for the Trailmaking-A and B have been estimated partly based on the same sample (Waggestad et al., 2025).

We have found higher prevalence of oral health problems among persons with cognitive impairments in both home-dwellers and nursing home residents (Asante et al., 2025).

Accelerometer data from the Trondheim 70+ study part has also shown that objectively measured daily physical activity is lower in higher age and lower in those receiving higher levels of care (Ustad et al., 2024). Older adults with mild cognitive impairment had lower physical performance and lower physical activity compared to those with normal cognitive function (Antonsen et al., 2021). There was a negative association between time spent physically active and the risk of a highly increasing trajectory of care needs (Ustad et al., 2025).

Supplemental table 7 outlines a number of studies based on the HUNT4 70+ cohort.

Supplemental table 3 – relative proportions of primary care diagnoses in women, HUNT4 Trondheim 70+

| **Women** | | | | | |
|---|---|---|---|---|---|
| **Diagnosis** | **70-74** | **75-79** | **80-84** | **85-89** | **90+** |
| D84 Oesophagus disease | 1.57 (0.89, 2.77) | 1.24 (0.58, 2.64) | 1.52 (0.58, 4.03) | - | 1.94 (0.75, 4.98) |
| H84 Presbyacusis | - | - | 1.73 (0.61, 4.88) | 1.55 (0.67, 3.6) | 0.63 (0.32, 1.23) |
| K74 Angina pectoris | - | 1.66 (0.54, 5.08) | 1.83 (0.73, 4.61) | 1.1 (0.52, 2.34) | 0.99 (0.53, 1.83) |
| K77 Heart failure | - | - | - | 0.8 (0.44, 1.46) | 0.83 (0.52, 1.34) |
| K78 Atrial fibrillation | 0.85 (0.5, 1.46) | 1.15 (0.65, 2.03) | 0.94 (0.58, 1.53) | 1.4 (0.92, 2.13) | 1.29 (0.87, 1.92) |
| K86 Arterial hypertension, uncomplicated | 0.81 (0.67, 0.98) | 0.87 (0.69, 1.09) | 1.01 (0.79, 1.29) | 0.92 (0.72, 1.18) | 0.85 (0.66, 1.11) |
| K87 Arterial hypertension, complicated | 1.18 (0.46, 3.03) | - | - | 1.48 (0.57, 3.85) | - |
| K90 Stroke | 1.39 (0.67, 2.87) | 0.73 (0.35, 1.52) | 1.3 (0.63, 2.68) | 1.59 (0.86, 2.93) | 1.5 (0.79, 2.83) |
| L88 Rheumatoid arthritis | 1.02 (0.57, 1.85) | 0.9 (0.43, 1.89) | - | 1.19 (0.51, 2.81) | 3.24 (1.08, 9.71) |
| L95 Osteoporosis | 1.3 (0.91, 1.85) | 0.86 (0.54, 1.37) | 1.07 (0.66, 1.74) | 0.94 (0.61, 1.43) | 1.11 (0.7, 1.76) |
| N17 Dizziness/vertigo | 1.26 (0.8, 1.99) | 1 (0.58, 1.73) | 1.14 (0.65, 1.99) | 0.6 (0.35, 1.04) | 0.97 (0.54, 1.73) |
| P06 Insomnia | 1.11 (0.78, 1.58) | 0.93 (0.57, 1.52) | 0.74 (0.46, 1.19) | 0.75 (0.45, 1.23) | 0.73 (0.42, 1.28) |
| P70 Dementia | 1.01 (0.51, 2) | 1.9 (0.98, 3.67) | 1.1 (0.66, 1.81) | 2.2 (1.47, 3.28) | 1.46 (0.99, 2.15) |
| P76 Depression | 1.18 (0.66, 2.12) | 1.01 (0.49, 2.11) | 1.41 (0.66, 3) | 1.34 (0.69, 2.59) | 1.34 (0.54, 3.3) |
| R95 COPD | 0.89 (0.59, 1.32) | 0.54 (0.32, 0.9) | 1.27 (0.68, 2.34) | 1.12 (0.57, 2.19) | - |
| R96 Astma | 0.59 (0.34, 1.02) | 1.85 (0.84, 4.07) | - | 1.46 (0.61, 3.51) | - |
| T86 Hypothyroidism | 0.46 (0.32, 0.67) | 0.8 (0.48, 1.33) | 0.92 (0.52, 1.62) | 0.74 (0.42, 1.32) | 0.98 (0.55, 1.75) |
| T90 Diabetes type 2 | 0.65 (0.46, 0.92) | 1.02 (0.66, 1.57) | 0.49 (0.3, 0.81) | 1.21 (0.75, 1.95) | - |
| U04 Urinary incontinence | 1.42 (0.8, 2.53) | 1.5 (0.78, 2.87) | 0.8 (0.42, 1.51) | 1.27 (0.74, 2.17) | 0.87 (0.52, 1.45) |

Relative proportions of diagnosed cases among the included participants compared to the local population.
Groups with <5 cases among participants are missing.

Supplemental table 4 – relative proportions of primary care diagnoses in men, HUNT4 Trondheim 70+

| Men | | | | | |
|---|---|---|---|---|---|
| **Diagnosis** | **70-74** | **75-79** | **80-84** | **85-89** | **90+** |
| D84 Oesophagus disease | 1.55 (0.77, 3.11) | - | - | - | - |
| H84 Presbyacusis | 1.56 (0.52, 4.62) | - | - | 2.36 (0.78, 7.14) | 2.9 (0.78, 10.76) |
| K74 Angina pectoris | 1.26 (0.65, 2.43) | 1.5 (0.57, 3.94) | - | - | - |
| K77 Heart failure | - | 1.2 (0.51, 2.85) | 0.9 (0.4, 2.01) | - | - |
| K78 Atrial fibrillation | 1.22 (0.84, 1.78) | 1.07 (0.68, 1.68) | 0.79 (0.49, 1.28) | 0.86 (0.53, 1.39) | 0.65 (0.35, 1.19) |
| K86 Arterial hypertension, uncomplicated | 0.99 (0.82, 1.21) | 0.94 (0.7, 1.28) | 1.26 (0.86, 1.84) | 1.12 (0.73, 1.74) | 1.29 (0.68, 2.46) |
| K87 Arterial hypertension, complicated | 0.79 (0.36, 1.71) | - | - | - | - |
| K90 Stroke | 1.38 (0.75, 2.52) | - | - | - | - |
| L88 Rheumatoid arthritis | 0.92 (0.4, 2.11) | - | - | - | - |
| L95 Osteoporosis | - | - | - | - | - |
| N17 Dizziness/vertigo | 0.96 (0.53, 1.74) | 2.6 (1.13, 5.95) | 1.12 (0.45, 2.77) | 1.46 (0.57, 3.71) | - |
| P06 Insomnia | 0.82 (0.49, 1.39) | 0.71 (0.35, 1.47) | 1.23 (0.52, 2.91) | - | - |
| P70 Dementia | 0.99 (0.45, 2.19) | 1.43 (0.66, 3.06) | 1.12 (0.53, 2.38) | 1.65 (0.83, 3.25) | 1.55 (0.61, 3.93) |
| P76 Depression | - | - | - | - | - |
| R95 COPD | 0.46 (0.31, 0.7) | 0.51 (0.28, 0.94) | 1.28 (0.63, 2.59) | - | - |
| R96 Astma | 0.74 (0.35, 1.57) | - | - | - | - |
| T86 Hypothyroidism | - | - | - | - | - |
| T90 Diabetes type 2 | 0.96 (0.72, 1.27) | 0.5 (0.34, 0.76) | 1.04 (0.61, 1.79) | 0.78 (0.4, 1.5) | - |
| U04 Urinary incontinence | - | - | - | - | - |

Relative proportions of diagnosed cases among the included participants compared to the local population.
Groups with <5 cases among participants are missing.

**Supplements for the main HUNT4 70+ cohort**

Supplemental table 5 Participant characteristics

| | Field station (N=8522) | Home visit (N=803) | Nursing home (N=631) | Overall (N=9956) |
|---|---|---|---|---|
| **Sex** | | | | |
| Female | 4454 (52.3%) | 545 (67.9%) | 421 (66.7%) | 5420 (54.4%) |
| Male | 4068 (47.7%) | 258 (32.1%) | 210 (33.3%) | 4536 (45.6%) |
| **Age stratum** | | | | |
| 70-79 | 6429 (75.4%) | 162 (20.2%) | 121 (19.2%) | 6712 (67.4%) |
| 80-89 | 1929 (22.6%) | 385 (47.9%) | 265 (42.0%) | 2579 (25.9%) |
| 90+ | 164 (1.9%) | 256 (31.9%) | 245 (38.8%) | 665 (6.7%) |
| **Education** | | | | |
| Primary | 2355 (27.6%) | 468 (58.3%) | 350 (55.5%) | 3173 (31.9%) |
| Secondary | 3871 (45.4%) | 197 (24.5%) | 149 (23.6%) | 4217 (42.4%) |
| College/University | 2261 (26.5%) | 57 (7.1%) | 44 (7.0%) | 2362 (23.7%) |
| **Smoking Status** | | | | |
| Never | 3204 (37.6%) | 214 (26.7%) | 130 (20.6%) | 3548 (35.6%) |
| Former | 4270 (50.1%) | 235 (29.3%) | 148 (23.5%) | 4653 (46.7%) |
| Current | 534 (6.3%) | 32 (4.0%) | 12 (1.9%) | 578 (5.8%) |
| Occasional | 28 (0.3%) | 0 (0%) | 0 (0%) | 28 (0.3%) |
| Former Occasional | 384 (4.5%) | 23 (2.9%) | 12 (1.9%) | 419 (4.2%) |
| **ApoE ε4 carrier** | | | | |
| No | 5960 (69.9%) | 560 (69.7%) | 355 (56.3%) | 6875 (69.1%) |
| Yes | 2501 (29.3%) | 223 (27.8%) | 246 (39.0%) | 2970 (29.8%) |
| **BMI ($kg/m^2$)** | 27.2 (4.3) | 27.1 (5.4) | 26.1 (4.8) | 27.2 (4.4) |
| **Body fat (%)** | 33.6 (8.5) | - | - | 33.6 (8.5) |
| **MUAC (cm)** | 30.1 (3.5) | 28.5 (4.2) | 27.9 (4.0) | 29.9 (3.6) |
| **Systolic BP (mmHg)** | 140.2 (19.8) | 135.7 (22.7) | 130.4 (22.2) | 139.4 (20.3) |
| **Grip Strength (kg)** | 33.7 (10.8) | 21.3 (7.8) | 18.1 (8.7) | 31.9 (11.5) |
| **SPPB** | 11 [9, 12] | 4 [2, 7] | 1 [0, 3] | 11 [8, 12] |
| **ROAG-J**[a] | | | | |
| Normal | - | 167 (20.8%) | 81 (12.8%) | - |
| ≥1 Moderate Problem | - | 371 (46.2%) | 245 (38.8%) | - |
| ≥1 Severe Problem | - | 161 (20.0%) | 100 (15.8%) | - |
| **MoCA** | 24 [21, 26] | 17 [12, 21] | 13 [9, 17] | 24 [20, 26] |
| **CERAD 10-word recall** | 5 [4, 6] | 4 [2, 6] | - | 5 [4, 6] |
| **Dementia assessment** | | | | |
| No cognitive impairment | 4698 (55.1%) | 114 (14.2%) | 10 (1.6%) | 4822 (48.4%) |
| aMCI | 2658 (31.2%) | 211 (26.3%) | 51 (8.1%) | 2920 (29.3%) |
| naMCI | 438 (5.1%) | 37 (4.6%) | 22 (3.5%) | 497 (5.0%) |
| Dementia | 618 (7.3%) | 390 (48.6%) | 523 (82.9%) | 1531 (15.4%) |
| Unclassified | 110 (1.3%) | 51 (6.4%) | 25 (4.0%) | 186 (1.9%) |

Continuous variables are presented as mean (SD), and discrete scores as median [IQR]. Percentages are of total N in column. Missing is not shown. BMI: Body Mass Index, MUAC: Middle Upper Arm Circumference, SPPB: Short Physical Performance Battery, ROAG-J: Revised Oral Assessment Guide – Jönköping, CERAD: Consortium to Establish a Registry for Alzheimer's Disease. aMCI: amnestic mild cognitive impairment (MCI), naMCI: non-amnestic MCI. [a]ROAG-J was only assessed at home visits (n=699) and nursing homes (n=426).

Supplemental table 6: Data availability

| Data | Field station (N=8522) | Home visit (N=803) | Nursing home (N=631) | Total (N=9956) |
|---|---|---|---|---|
| **Questionnaire** | | | | |
| Education | 8487 ( 99.6 %) | 722 ( 89.9 %) | 543 ( 86.1 %) | 9752 ( 98 %) |
| Smoking status | 8420 ( 98.8 %) | 504 ( 62.8 %) | 302 ( 47.9 %) | 9226 ( 92.7 %) |
| Weight loss | 7805 ( 91.6 %) | 433 ( 53.9 %) | 209 ( 33.1 %) | 8447 ( 84.8 %) |
| Exhaustion | 7147 ( 83.9 %) | 455 ( 56.7 %) | 224 ( 35.5 %) | 7826 ( 78.6 %) |
| HADS | 7002 ( 82.2 %) | 389 ( 48.4 %) | 163 ( 25.8 %) | 7554 ( 75.9 %) |
| **Clinical examinations** | | | | |
| BMI | 8458 ( 99.2 %) | 688 ( 85.7 %) | 312 ( 49.4 %) | 9458 ( 95 %) |
| Body Composition | 7565 ( 88.8 %) | - | - | 7565 ( 76 %) |
| MUAC | 7364 ( 86.4 %) | 791 ( 98.5 %) | 527 ( 83.5 %) | 8682 ( 87.2 %) |
| Systolic BP | 8492 ( 99.6 %) | 775 ( 96.5 %) | 524 ( 83 %) | 9791 ( 98.3 %) |
| Gait Speed | 8206 ( 96.3 %) | 625 ( 77.8 %) | 311 ( 49.3 %) | 9142 ( 91.8 %) |
| Grip Strength | 8091 ( 94.9 %) | 765 ( 95.3 %) | 481 ( 76.2 %) | 9337 ( 93.8 %) |
| SPPB | 7874 ( 92.4 %) | 703 ( 87.5 %) | 490 ( 77.7 %) | 9067 ( 91.1 %) |
| 1-week Accelerometer recording | 4512 ( 52.9 %) | 244 ( 30.4 %) | 107 ( 17 %) | 4863 ( 48.8 %) |
| ROAG-J | - | 699 ( 87 %) | 426 ( 67.5 %) | 1125 ( 11.3 %) |
| MoCA | 8116 ( 95.2 %) | 683 ( 85.1 %) | 109 ( 17.3 %) | 8908 ( 89.5 %) |
| CERAD 10-word recall | 5338 ( 62.6 %) | 101 ( 12.6 %) | - | 5439 ( 54.6 %) |
| SIB-8 | - | - | 356 ( 56.4 %) | 356 ( 3.6 %) |
| Dementia assessment | 8417 ( 98.8 %) | 752 ( 93.6 %) | 606 ( 96 %) | 9775 ( 98.2 %) |
| **Lab data (examples)** | | | | |
| Cholesterol | 8433 ( 99 %) | 637 ( 79.3 %) | 469 ( 74.3 %) | 9539 ( 95.8 %) |
| pTau-217 | 8030 ( 94.2 %) | 531 ( 66.1 %) | 387 ( 61.3 %) | 8948 ( 89.9 %) |
| NfL | 7888 ( 92.6 %) | 518 ( 64.5 %) | 383 ( 60.7 %) | 8789 ( 88.3 %) |
| Genetics | 8501 ( 99.8 %) | 785 ( 97.8 %) | 603 ( 95.6 %) | 9889 ( 99.3 %) |
| **Biosamples** | | | | |
| Serum | 8394 ( 98.5 %) | 618 ( 77 %) | 456 ( 72.3 %) | 9468 ( 95.1 %) |
| Plasma | 8128 ( 95.4 %) | 538 ( 67 %) | 397 ( 62.9 %) | 9063 ( 91 %) |
| Urine | 4251 ( 49.9 %) | 104 ( 13 %) | 36 ( 5.7 %) | 4391 ( 44.1 %) |
| Faeces | 3016 ( 35.4 %) | 26 ( 3.2 %) | 9 ( 1.4 %) | 3051 ( 30.6 %) |
| Saliva | 2939 ( 34.5 %) | 85 ( 10.6 %) | 20 ( 3.2 %) | 3044 ( 30.6 %) |
| HUNT3 Serum | 7432 ( 87.2 %) | 553 ( 68.9 %) | 400 ( 63.4 %) | 8385 ( 84.2 %) |
| HUNT3 Plasma | 6593 ( 77.4 %) | 532 ( 66.3 %) | 381 ( 60.4 %) | 7506 ( 75.4 %) |
| HUNT2 Serum | 7487 ( 87.9 %) | 704 ( 87.7 %) | 529 ( 83.8 %) | 8720 ( 87.6 %) |

HADS: Hospital Anxiety and Depression Scale, BMI: Body Mass Index, MUAC: Middle Upper Arm Circumference, SPPB: Short Physical Performance Battery, ROAG-J: Revised Oral Assessment Guide – Jönköping, MoCA: Montreal Cognitive Assessment, CERAD: Consortium to Establish a Registry for Alzheimer's Disease. SIB-8: Severe Impairment Battery-8. pTau-217: plasma phosphorylated Tau-217, NfL, plasma neurofilament light.

Supplemental table 7 – relative proportions of primary care diagnoses in women,
**The main HUNT4 70+ cohort**

| Women | | | | | |
|---|---|---|---|---|---|
| **Diagnosis** | **70-74** | **75-79** | **80-84** | **85-89** | **90+** |
| D84 Oesophagus disease | 0.92 (0.68, 1.24) | 0.71 (0.52, 0.97) | 1.05 (0.75, 1.47) | 1.11 (0.71, 1.73) | 1.36 (0.88, 2.1) |
| H84 Presbyacusis | 0.78 (0.45, 1.35) | 1.32 (0.81, 2.15) | 1 (0.66, 1.51) | 1.13 (0.73, 1.75) | 1.59 (1, 2.51) |
| H86 Deafness | 1.93 (1, 3.74) | 1.38 (0.85, 2.25) | 1.03 (0.64, 1.66) | 1.52 (0.85, 2.72) | 0.99 (0.62, 1.58) |
| K74 Angina pectoris | 0.94 (0.62, 1.42) | 0.74 (0.53, 1.03) | 1.07 (0.74, 1.54) | 0.65 (0.48, 0.88) | 1.01 (0.78, 1.32) |
| K75 Myocardial infarction | 1.39 (0.5, 3.9) | 0.73 (0.37, 1.43) | 0.98 (0.47, 2.03) | 0.78 (0.43, 1.44) | 0.72 (0.43, 1.21) |
| K76 Coronary artery disease | 1.14 (0.75, 1.72) | 0.76 (0.54, 1.05) | 0.73 (0.49, 1.07) | 0.98 (0.64, 1.5) | 0.86 (0.59, 1.25) |
| K77 Heart failure | 0.39 (0.25, 0.6) | 0.71 (0.53, 0.94) | 0.71 (0.56, 0.9) | 1.17 (0.93, 1.46) | 0.95 (0.79, 1.14) |
| K78 Atrial fibrillation | 0.94 (0.74, 1.21) | 0.75 (0.63, 0.89) | 0.87 (0.73, 1.03) | 0.96 (0.81, 1.13) | 0.9 (0.76, 1.07) |
| K85 Elevated Blood Pressure | 1.25 (0.86, 1.82) | 1.02 (0.74, 1.41) | 1.42 (0.97, 2.09) | 0.95 (0.6, 1.49) | 1.31 (0.81, 2.12) |
| K86 Arterial hypertension, uncomplicated | 0.96 (0.88, 1.05) | 1.03 (0.95, 1.11) | 0.93 (0.85, 1.02) | 0.87 (0.79, 0.96) | 1.03 (0.92, 1.16) |
| K87 Arterial hypertension, complicated | 0.71 (0.49, 1.03) | 0.77 (0.57, 1.05) | 1.02 (0.7, 1.47) | 1.08 (0.76, 1.55) | 1.05 (0.74, 1.47) |
| K89 TIA | 0.65 (0.41, 1.03) | 0.84 (0.58, 1.23) | 0.89 (0.61, 1.3) | 1.28 (0.81, 2.02) | 1.27 (0.89, 1.82) |
| K90 Stroke | 1.36 (0.93, 1.99) | 0.94 (0.71, 1.24) | 0.95 (0.73, 1.24) | 0.89 (0.69, 1.15) | 0.93 (0.73, 1.18) |
| K91 Cerebrovascular disease | 0.66 (0.39, 1.12) | 0.93 (0.63, 1.37) | 0.75 (0.5, 1.13) | 0.89 (0.56, 1.4) | 1.59 (0.96, 2.65) |
| L18 Myalgia | 0.78 (0.6, 1.01) | 0.86 (0.66, 1.11) | 1.26 (0.88, 1.79) | 0.7 (0.48, 1.02) | 1.35 (0.79, 2.29) |
| L86 Radiating low back pain | 0.87 (0.65, 1.17) | 1.14 (0.81, 1.6) | 1.04 (0.69, 1.56) | 1.22 (0.71, 2.08) | 1.32 (0.69, 2.52) |
| L88 Rheumatoid arthritis | 0.79 (0.6, 1.05) | 0.92 (0.7, 1.2) | 0.75 (0.56, 1.01) | 0.89 (0.6, 1.31) | 0.99 (0.63, 1.56) |
| L95 Osteoporosis | 1.42 (1.14, 1.76) | 1.53 (1.28, 1.84) | 1.26 (1.04, 1.53) | 1.11 (0.91, 1.36) | 1.46 (1.17, 1.83) |
| N17 Dizziness/vertigo | 0.94 (0.73, 1.21) | 0.85 (0.69, 1.05) | 1.03 (0.82, 1.3) | 1.18 (0.9, 1.56) | 0.79 (0.6, 1.06) |
| P06 Insomnia | 0.97 (0.81, 1.15) | 0.9 (0.77, 1.05) | 1.03 (0.87, 1.22) | 0.81 (0.67, 0.97) | 1.01 (0.81, 1.27) |
| P20 Memory disturbance | 0.69 (0.42, 1.12) | 0.55 (0.41, 0.74) | 1.09 (0.8, 1.47) | 1.14 (0.83, 1.57) | 1.07 (0.78, 1.47) |
| P70 Dementia | 0.98 (0.66, 1.46) | 0.74 (0.59, 0.92) | 0.88 (0.72, 1.08) | 1.1 (0.92, 1.31) | 0.9 (0.75, 1.07) |
| P74 Anxiety | 0.74 (0.52, 1.04) | 0.69 (0.5, 0.97) | 0.72 (0.48, 1.07) | 0.87 (0.58, 1.31) | 1.02 (0.68, 1.54) |
| P76 Depression | 0.64 (0.5, 0.82) | 0.62 (0.5, 0.77) | 0.86 (0.68, 1.09) | 0.75 (0.58, 0.97) | 1.16 (0.87, 1.55) |
| R95 COPD | 0.75 (0.62, 0.92) | 0.9 (0.75, 1.07) | 0.62 (0.5, 0.76) | 1.06 (0.8, 1.41) | 1.01 (0.72, 1.42) |
| R96 Asthma | 1.44 (1.07, 1.93) | 0.91 (0.7, 1.18) | 0.93 (0.66, 1.31) | 1.01 (0.7, 1.46) | 1.16 (0.71, 1.9) |
| T86 Hypothyroidism | 1.22 (1.01, 1.49) | 0.83 (0.7, 0.99) | 1.24 (1, 1.55) | 0.84 (0.65, 1.08) | 0.84 (0.64, 1.09) |
| T89 Diabetes type 1 | 0.7 (0.41, 1.21) | 0.67 (0.45, 1.02) | 1.12 (0.67, 1.87) | 1.08 (0.57, 2.06) | 0.73 (0.42, 1.27) |
| T90 Diabetes type 2 | 0.82 (0.7, 0.95) | 0.75 (0.65, 0.87) | 0.83 (0.71, 0.98) | 0.98 (0.81, 1.19) | 1.02 (0.8, 1.32) |
| T92 Arthitis hyperurecemia | 1.14 (0.68, 1.93) | 0.47 (0.31, 0.7) | 1.35 (0.84, 2.16) | 1.33 (0.85, 2.11) | 1.15 (0.71, 1.84) |
| U04 Urinary incontinence | 0.99 (0.75, 1.31) | 0.74 (0.6, 0.92) | 1.1 (0.87, 1.4) | 0.75 (0.61, 0.93) | 0.97 (0.78, 1.2) |

Relative proportions of diagnosed cases among the included participants compared to the invited population.
Groups with <5 cases among participants are missing.

Supplemental table 8 – relative proportions of primary care diagnoses in men,
**The main HUNT4 70+ cohort**

| Men | | | | | |
|---|---|---|---|---|---|
| **Diagnosis** | **70-74** | **75-79** | **80-84** | **85-89** | **90+** |
| D84 Oesophagus disease | 0.75 (0.53, 1.05) | 1.26 (0.9, 1.75) | 0.74 (0.47, 1.15) | 0.8 (0.47, 1.34) | 0.79 (0.41, 1.52) |
| H84 Presbyacusis | 2.04 (1.04, 4) | 1.1 (0.73, 1.65) | 1.27 (0.75, 2.16) | 0.94 (0.55, 1.61) | 1.38 (0.73, 2.58) |
| H86 Deafness | 1.08 (0.7, 1.67) | 1.43 (0.93, 2.2) | 1.2 (0.7, 2.03) | 0.97 (0.55, 1.73) | 1.06 (0.52, 2.14) |
| K74 Angina pectoris | 0.87 (0.65, 1.17) | 0.93 (0.7, 1.23) | 1.12 (0.82, 1.52) | 0.81 (0.56, 1.17) | 1.01 (0.73, 1.39) |
| K75 Myocardial infarction | 0.76 (0.44, 1.31) | - | 0.95 (0.5, 1.79) | - | 0.73 (0.37, 1.44) |
| K76 Coronary artery disease | 0.92 (0.73, 1.16) | 0.82 (0.67, 1) | 1.06 (0.81, 1.39) | 0.74 (0.51, 1.07) | 0.61 (0.42, 0.87) |
| K77 Heart failure | 0.6 (0.42, 0.85) | 0.88 (0.68, 1.14) | 0.72 (0.57, 0.92) | 0.75 (0.58, 0.98) | 0.87 (0.68, 1.13) |
| K78 Atrial fibrillation | 1.04 (0.87, 1.24) | 0.83 (0.73, 0.95) | 0.76 (0.66, 0.87) | 0.89 (0.75, 1.05) | 0.95 (0.78, 1.15) |
| K85 Elevated Blood Pressure | 0.62 (0.45, 0.84) | 1.05 (0.75, 1.48) | 0.83 (0.55, 1.25) | 1.63 (0.94, 2.82) | 0.88 (0.39, 2) |
| K86 Arterial hypertension, uncomplicated | 0.87 (0.79, 0.95) | 1.08 (0.98, 1.18) | 1.15 (1.01, 1.31) | 1.18 (1, 1.4) | 0.93 (0.75, 1.16) |
| K87 Arterial hypertension, complicated | 0.73 (0.5, 1.06) | 0.81 (0.61, 1.09) | 0.93 (0.66, 1.29) | 1.28 (0.75, 2.17) | 0.83 (0.49, 1.41) |
| K89 TIA | 1.14 (0.67, 1.96) | 0.91 (0.59, 1.39) | 1.02 (0.63, 1.63) | 0.76 (0.43, 1.34) | 1.09 (0.59, 2.01) |
| K90 Stroke | 0.74 (0.56, 0.97) | 0.74 (0.59, 0.92) | 0.61 (0.48, 0.78) | 0.91 (0.68, 1.21) | 0.97 (0.7, 1.36) |
| K91 Cerebrovascular disease | 1.04 (0.68, 1.59) | 1.21 (0.85, 1.73) | 0.56 (0.38, 0.82) | 0.66 (0.41, 1.05) | 1.48 (0.79, 2.78) |
| L18 Myalgia | 1.1 (0.67, 1.8) | 0.89 (0.58, 1.37) | 1.39 (0.75, 2.58) | 1.25 (0.58, 2.7) | 1.59 (0.58, 4.33) |
| L86 Radiating low back pain | 1 (0.69, 1.45) | 1.18 (0.77, 1.82) | 0.69 (0.44, 1.07) | 1.02 (0.51, 2.03) | 1.32 (0.49, 3.61) |
| L88 Rheumatoid arthritis | 0.84 (0.59, 1.2) | 0.56 (0.39, 0.81) | 0.9 (0.57, 1.42) | 0.91 (0.49, 1.7) | 1.24 (0.55, 2.79) |
| L95 Osteoporosis | 1.14 (0.59, 2.21) | 0.66 (0.4, 1.08) | 0.98 (0.56, 1.7) | 1.36 (0.66, 2.77) | - |
| N17 Dizziness/vertigo | 0.94 (0.68, 1.31) | 0.93 (0.71, 1.22) | 1.31 (0.93, 1.83) | 1.27 (0.86, 1.88) | 0.91 (0.58, 1.43) |
| P06 Insomnia | 0.92 (0.71, 1.2) | 1.01 (0.79, 1.29) | 0.93 (0.72, 1.21) | 0.69 (0.53, 0.91) | 0.89 (0.66, 1.21) |
| P20 Memory disturbance | 1.08 (0.69, 1.67) | 0.55 (0.4, 0.76) | 0.95 (0.69, 1.31) | 1.34 (0.87, 2.05) | 0.88 (0.59, 1.31) |
| P70 Dementia | 1.18 (0.68, 2.04) | 0.58 (0.44, 0.75) | 0.9 (0.71, 1.14) | 0.82 (0.64, 1.05) | 1.23 (0.91, 1.67) |
| P74 Anxiety | 0.68 (0.41, 1.13) | 0.68 (0.35, 1.3) | - | 1.29 (0.56, 2.99) | 0.61 (0.31, 1.2) |
| P76 Depressioin | 0.91 (0.6, 1.39) | 0.79 (0.56, 1.11) | 0.72 (0.48, 1.08) | 0.72 (0.44, 1.18) | 0.83 (0.5, 1.37) |
| R95 COPD | 0.98 (0.8, 1.2) | 0.77 (0.65, 0.92) | 0.73 (0.59, 0.9) | 0.63 (0.48, 0.81) | 1.38 (0.95, 2.02) |
| R96 Astma | 1.24 (0.86, 1.79) | 1.19 (0.83, 1.72) | 0.79 (0.53, 1.18) | 0.65 (0.37, 1.13) | 1.43 (0.66, 3.1) |
| T86 Hypothyroidism | 1.11 (0.76, 1.61) | 0.92 (0.66, 1.28) | 1.3 (0.84, 2) | 0.91 (0.49, 1.7) | 0.71 (0.44, 1.12) |
| T89 Diabetes type 1 | 0.75 (0.49, 1.17) | 0.89 (0.57, 1.4) | 1.39 (0.85, 2.3) | 0.71 (0.4, 1.26) | 1.11 (0.51, 2.41) |
| T90 Diabetes type 2 | 1 (0.87, 1.14) | 0.85 (0.75, 0.96) | 0.97 (0.82, 1.15) | 0.86 (0.67, 1.1) | 1.1 (0.8, 1.5) |
| T92 Arthitis hyperurecemia | 1.04 (0.75, 1.45) | 1.13 (0.86, 1.49) | 0.67 (0.5, 0.91) | 1.04 (0.68, 1.6) | 1.38 (0.84, 2.28) |
| U04 Urinary incontinence | 0.58 (0.39, 0.88) | 1.08 (0.77, 1.52) | 1.17 (0.79, 1.73) | 0.85 (0.57, 1.25) | 0.77 (0.51, 1.17) |

Relative proportions of diagnosed cases among the included participants compared to the invited population.
Groups with <5 cases among participants are missing.

Supplemental table 9: Overview of published studies from HUNT4 70+

| | |
|---|---|
| **Dementia risk and association** | |
| Blood pressure | (Lerfald et al., 2024; Lerfald et al., 2023; Selbaek et al., 2022) |
| Body mass index and waist circumference | (Zotcheva et al., 2025) |
| Depression and anxiety | (Aunsmo et al., 2024) |
| Education | (Mekonnen, Skirbekk, Håberg, et al., 2025) |
| Health behavior and physical activity | (Angelsen et al., 2024) |
| Hearing | (Moradi et al., 2022, 2023; Myrstad et al., 2023; Myrstad et al., 2025) |
| Infection | (Kelsey et al., 2025) |
| Marital status | (Skirbekk et al., 2023) |
| Nutritional factors | (Abbel et al., 2023; Asante et al., 2023) |
| Number of children | (Mekonnen, Skirbekk, Zotcheva, et al., 2025) |
| Occupation and retirement | (Edwin et al., 2024; Zotcheva, Strand, et al., 2023) |
| Physical activity | (Zotcheva, Bratsberg, et al., 2023) |
| Physical performance | (Antonsen et al., 2021; Sverdrup et al., 2021) |
| Sex | (Engedal et al., 2021; Wedatilake et al., 2024) |
| Sleep | (Selbaek-Tungevåg et al., 2023) |
| Social isolation and social media use | (Ibsen, Zotcheva, Bergh, Gerritsen, Livingston, Lurås, Mamelund, Mork Rokstad, et al., 2025) |
| **Disability** | |
| Disability-free life expectancy | (Storeng et al., 2021) |
| **Health-care utilization** | |
| Physical activity across levels of care | (Ustad et al., 2024) |
| Care trajectories related to physical activity | (Ustad et al., 2025) |
| Health care utilization and medication use related to COVID-19 | (Ibsen et al., 2024; Ibsen, Zotcheva, Bergh, Gerritsen, Livingston, Lurås, Mamelund, Rokstad, et al., 2025) |
| **Prevalence and incidence** | |
| Clinical dementia | (Gjøra et al., 2021; Gjøra et al., 2024; Gjøra et al., 2023; Molvik et al., 2025) |
| Alzheimer's disease biomarkers | (Sunde et al., 2025) |
| Frailty | (Kyrdalen et al., 2024) |
| Malnutrition | (Kolberg et al., 2023) |
| Oral health problems | (Asante et al., 2025; Cetrelli et al., 2025) |
| **Validation and reference of scales** | |
| CERAD 10-word list task | (Wagle et al., 2022) |
| Hospital Anxiety and Depression Scale (HADS) | (Sivertsen et al., 2023) |
| Mini-Mental State Examination (MMSE-NR3) | (Engedal et al., 2023) |
| Montreal Cognitive Assessment (MoCA) | (Engedal et al., 2022) |
| Trail-making A and B | (Waggestad et al., 2025) |
| Short Physical Performance Battery (SPPB) and Grip Strength | (Melsæter et al., 2022) |
